# CoViD-19 Epidemic in India and Projections: Is Relief in Sight?

**DOI:** 10.1101/2020.05.08.20096008

**Authors:** Abhaya Indrayan, Shubham Shukla

## Abstract

**Background:** Projection of cases and deaths in an epidemic such as CoViD-19 is hazardous and the early projections were way-off the actual pattern. However, we now have actual data for more than 50 consecutive days in India that can be effectively used for projection.

**Methods:** We closely track the trend and use the same pattern for projection. We call this Empirical Model. We also fit a Theoretical Model based on a Gamma function on the pattern of some of the previous epidemics.

**Results:** The Empirical Model predicts the peak around the fourth week of May and the near end of the epidemic by the end of June 2020. The maximum number of active cases is likely to be nearly 75,000 during the second week of June. This would mean a peak demand of nearly 15,000 beds and nearly 4000 ventilators. The case-fatality based on those who have reached an outcome was nearly 10% in the first week of May and is likely to remain at this level for some time. Theoretical Model projected a peak of nearly 2500 new cases per day in the second week of May that seems to have been already breached. This model predicts the near end of the epidemic by the middle of July 2020.

**Conclusion:** With the current trend, the end of the epidemic is in sight with relatively mild consequences in India compared with most other countries.

## Introduction

A large number of models are floating around regarding the shape of the pandemic of Corona Virus Disease-2019 (CoViD-19) for various countries. Among them, the Singapore models^1^ and CDDEP models^2^ are popular. These models are available for several countries, including India. Some India-specific studies are also available based on early data. A study by Ranjan^3^ estimated a final epidemic size of 13,000 cases and expected that India will enter equilibrium by the end of May 2020. This study considered three models: (i) exponential model, (ii) logistic model, and (iii) SIR model. Menon^4^ described four studies based on SIER (Susceptible, Infected, Exposed, and Recovered) model. These are: (i) ICMR study that suggested 100 cases at the peak for every 10,000 people under optimistic scenario, amounting to 1.35 crore infected, (ii) Michigan study that predicted 16 cases per 100,000 population in the absence of any intervention, (iii) Hopkins study, which originates from IndiaSIM model, which predicted 1 crore to 2.5 crore affected in India at its peak, and (iv) Cambridge model that considered 21-day lockdown and expected a decline immediately thereafter. The author rightly said that none is believable. In any case, these were early efforts when data availability was scanty.

Among recent India-specific studies, Schueller et al.^5^ estimated a basic reproduction number (*R*_0_) of 2.66 under no intervention, 2.00 under moderate lockdown, and 1.5 under hard lockdown. They did not predict the number of cases. Ghosal et al.^6^ used linear regression to predict an average of 211 deaths per day in week 5 and 467 in week 6 of the epidemic. They did not go beyond this period and did not predict the number of cases.

India-specific articles cited in the preceding paragraph are pre-prints and have not undergone peer review. Yet, these attempts show that predicting the course of the epidemic can be hazardous. Now that since the data are available for more than 50 consecutive days starting from the day when the cumulative cases were more than 100, more plausible prediction (we are calling it ‘projection’) can be made. This communication reports our projections for the cases, the recoveries, and the deaths – and also the case-fatality.

## Material and Methods

The data on actual number of cumulative cases, recoveries, and deaths was taken from *COVID19India.org* for each day since the detection of the first case on 30th January 2020. However, the initial occurrence was erratic and our study is based on the data from 14th March 2020 when the cumulative number of cases was 100. New cases, new recoveries, and new deaths were calculated for each day till 5^th^ May 2020. The case-fatality was based on the deaths to recovery ratio as these are the cases whose outcome is known – others are still active whose recovery or death is not known. Death rate based on cumulative deaths and cumulative cases, as is generally reported, does not give a correct picture of the case-fatality.

The rate of increase of new cases, new recoveries, and new deaths per day was empirically studied for trend in terms of percentages, without using any mathematical model. This rate defines the acceleration or deceleration. This trend was extended to the period till the end of May in the first instance and then till the end of June when it was found that the projection worked fairly well for the last 15 days from 20^th^ April 2020 to 5^th^ May 2020. We call it an Empirical Model.

Many epidemics in the past such as of SARS^7^, MERS^8,9^, and HIV^10^ had skewness to the right, and it looks plausible also that the rise will be sharp and decline slow. Liu et al.^11^ found a similar pattern with CoViD epidemic. Accordingly, we investigated Gamma function that has this property. The function is as follows:

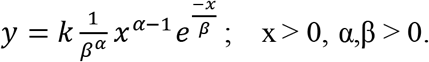

The method of maximum likelihood was used to estimate the parameters α and β^12^. We call this Theoretical Model.

Projections were done with both the models.

## Results

The situation on 5^th^ May 2020 is shown in Table 1. Nearly 32.1% reached to the outcome and the death rate based on the cumulative numbers was 3.4%. However, factually, 1693 died and 14142 recovered by this time – thus the death to recovery ratio is nearly 8:1. This really means 1 death out of 9 whose outcome is known, and gives case-fatality of nearly 11%. On 5 ^th^ May 2020, this rate was 127/1424 or nearly 8.9%. The case-fatality was nearly 20% in the early phase but showed continuous decline to settle at nearly 10% for the past several days.

**Table 1.** Actual situation of CoViD cases and deaths on 5^th^ May 2020

|  | Cases | Outcome |  |  |
| --- | --- | --- | --- | --- |
|  |  | Recoveries | Deaths | Total with outcome |
| Cumulative | 49400 | 14142 | 1693 | 15835 (32.1%) |
| New | 2966 | 1297 | 127 | 1424 |

### Empirical Model

Increase in the number of cases, which was nearly 20% per day at one point in time, has shown consistent pattern of decrease in acceleration of nearly 0.5% everyday for the past several days, with an exception of some occurrences in the first week of May and was 6.4% on 5^th^ May 2020. This decline in the rate of increase could be the direct result of social distancing and lockdown. We expect this decline will continue for quite some time except towards the end, after reaching to 0.1%. The rate of decline of increase is expected to slow down at this level because some cases will continue to crop up. When this is used for projection, the cumulative number of cases is likely to be 1,27,000 by the end of May and 1,75,000 by the end of June 2020. With this trend, the number of new cases is likely to peak to nearly 3300 per day during the fourth week of May after which it will decline, and the expected shape of the epidemic curve is shown in Figure 1. At this rate, most of the epidemic is likely to end by the fourth week of June.

**Figure 1.**
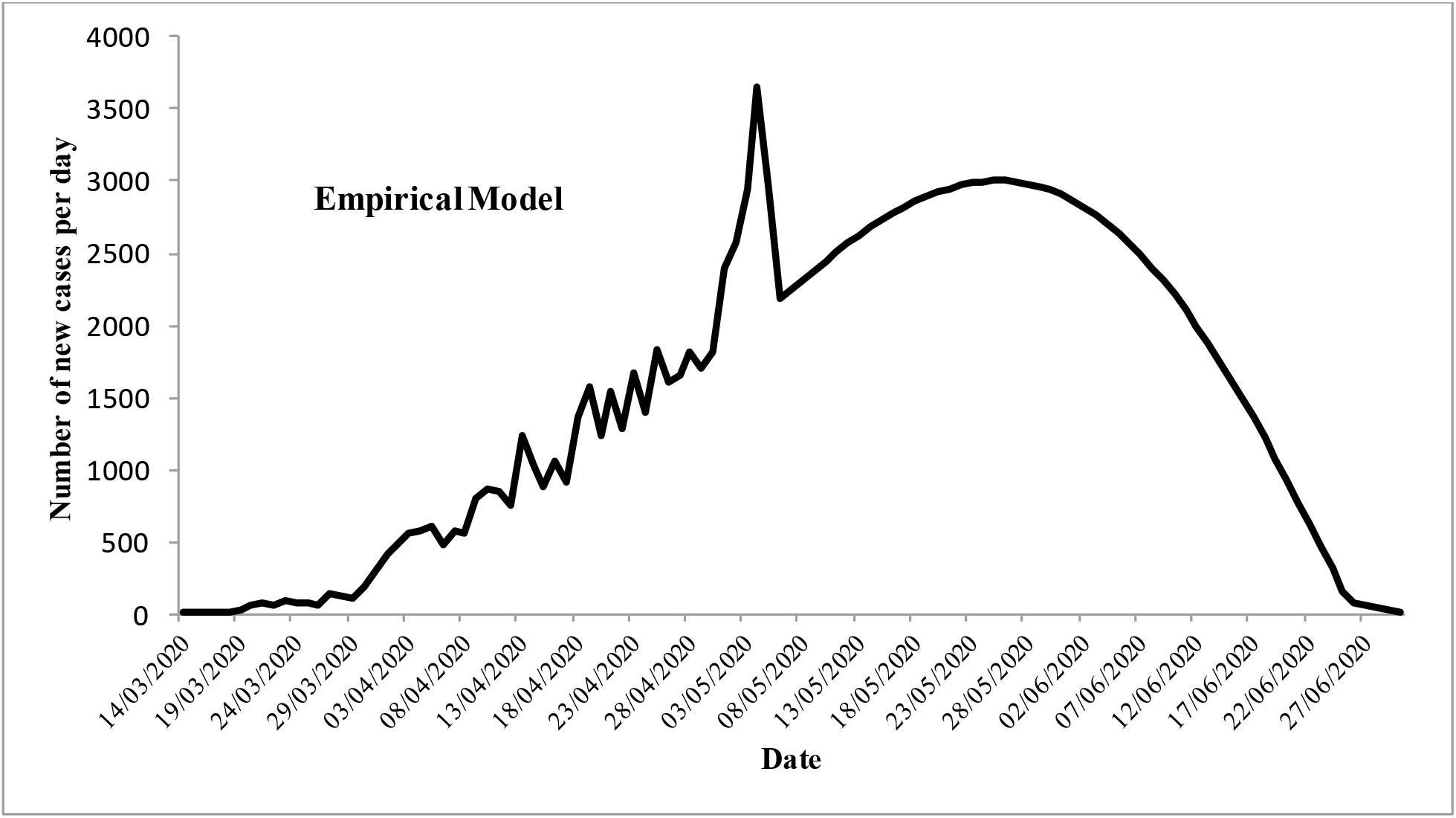
Shape of the epidemic by Empirical Model

Similar to the new cases, the cumulative recoveries have also shown an increasing pattern at the rate of 0.6% each day to reach from 10.3% on 10^th^ April to 28.6% on 5^th^ May 2020, with a slightly decreasing trend of 0.5% in the last 10 days. At this rate, the cumulative recoveries are expected to be 41% by the end of May and 56% by the end of June 2020. This will mean nearly 100,000 cases recovered by the end of June 2020.

Case fatality on the basis of those whose outcome is known also declined from nearly 25% in the early phase of the epidemic to now nearly 10% in the first week of May. This is stable at this level (Figure 2).

**Figure 2.**
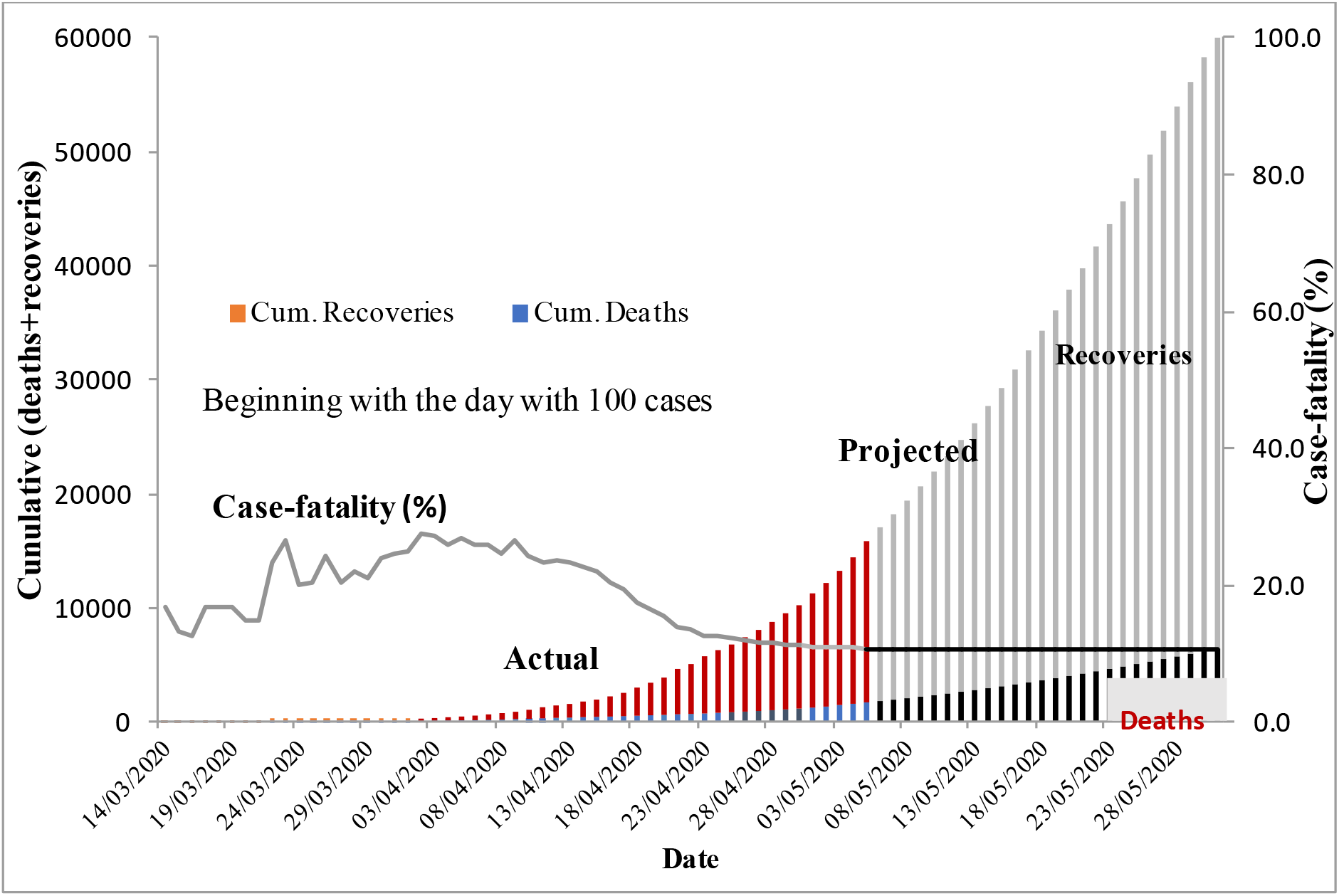
Recoveries, deaths and the case-fatality

Active cases are those who are still undergoing treatment and whose outcome either in terms of recovery or death is not known yet. These define the load on medical care services. This model indicates that the active cases are likely to be the highest at 75,000 during the 2nd week of June 2020. If 20% of these require hospitalization and 10% ventilator services, the peak requirement of beds is expected to be 15,000, each for nearly 10 days, and of ventilators nearly 7500.

### Theoretical Model

This model is based on Gamma function and assumes that the decline in the number of new cases will be slower that the rise during the first half of the epidemic. Thus, the shape is slightly skewed to the right.

The best estimate of the slope parameter and scale parameter of the Gamma function are α = 6.5 and β = 9.5, respectively. The shape of the predicted epidemic curve is shown in Figure 3. This indicates that nearly 99% of the epidemic will be over by the middle of July. The peak will occur around second week of May and the maximum number of new cases at that time will be nearly 2500 per day. As we go to the press, the projection by this model is already too low.

**Figure 3.**
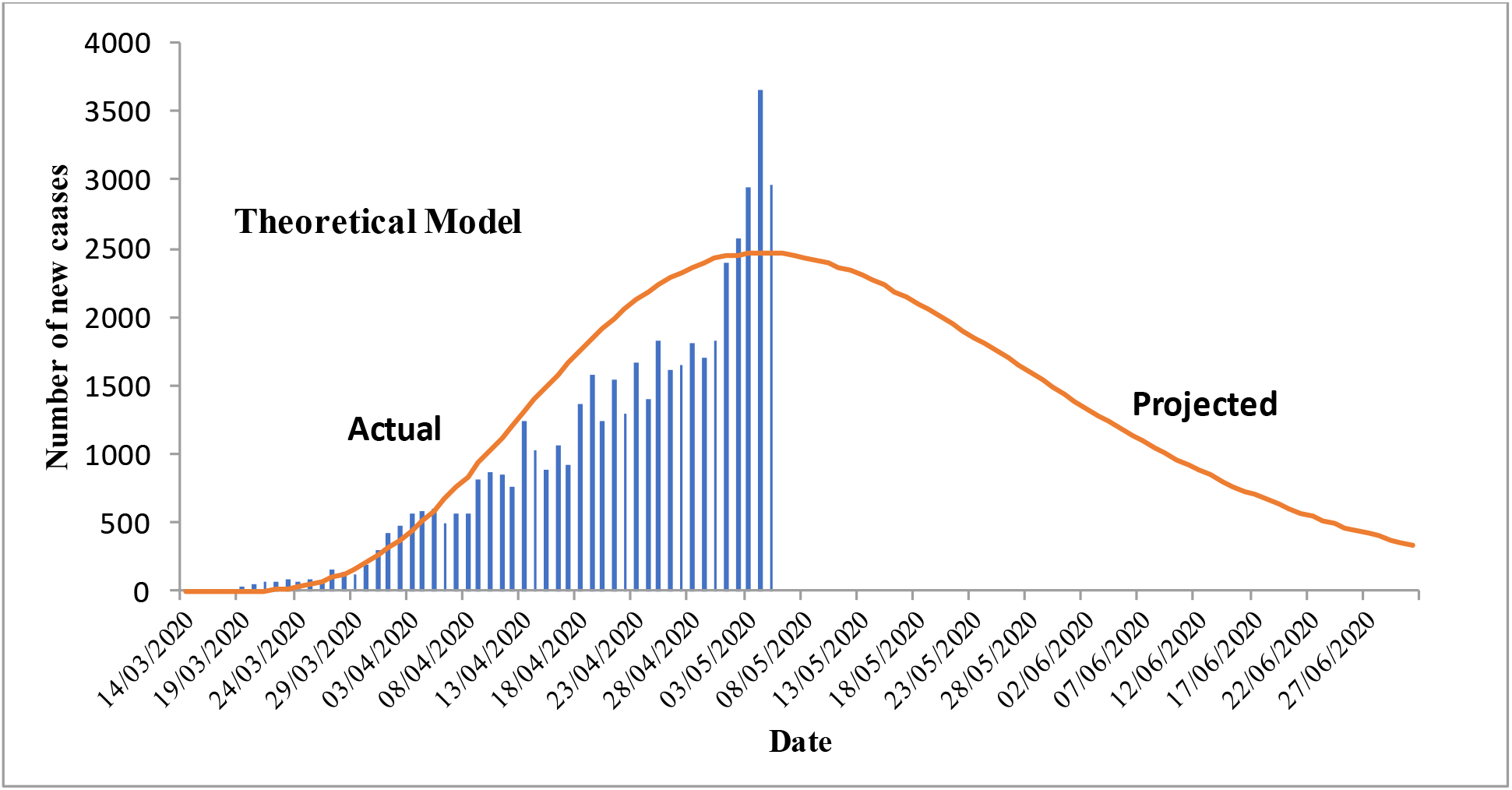
The theoretical model based on Gamma function

## Discussion

Our Empirical Model is based on the trend of the rate of occurrence of new cases relative to the cumulative cases on the previous day. This predicts a rapid decline after the peak of nearly 3300 per day around fourth week of May. Against this, the Theoretical Model based on Gamma function has slow decline and predicts a peak of 2500 new cases around the second week of May. The Empirical Model looks more plausible, first because it is based on the pattern actually observed, and second because the heat and ultra-violet rays in sunshine during the month of May and June in large part of the country can substantially help in reducing the transmission^13^.

Our projection seem more plausible. Chaterjee et al.^14^ estimated 241,974 total infections when all non-pharmacological interventions are adopted. Pandey et al.^15^ estimated 5300 new cases on 13th April with SEIR model and 6135 with regression model. Tiwari et al.^16^ predicted control of the CoViD epidemic by the end of May 2020 and the total cases around 68,978 at that time. The prediction in the first two papers quoted above is too high and the prediction in the third looks too low as the cumulative number has already exceeded 45,000 on 5^th^ May 2020.

Two important findings emerge that have not found much traction in public discourse. First, a large percentage of cases are recovering despite no treatment. This underscores the importance of our in-built immunity, either due to vegetarian diet in a large segment of population or due to highly prevalent BCG vaccination^17^.The second is that the case-fatality is not less than 5% as is generally considered^18^. When the cases with known outcomes are considered, the case-fatality is nearly 10%. This may eventually decline as our medical care system learns to manage these cases and new strategies such as convalescent plasma therapy^19^ and drugs such as Remedesivir^20^ succeed.

Our models, particularly the Empirical Model, are based on the trend and not any conventional epidemic modeling such as SEIR or time series. We are also not using the reproduction number (*R*_0_) that is commonly used for infectious disease modeling. This may look like a limitation but the projection is based on the trend seen for more than 50 consecutive days. This is substantial to study the trend and projection that predicts much less suffering and loss of life in India compared with most European countries and the U.S. We hope that we have been able to capture the salient feature of this trend and the projection will be largely true that could bring the relief in sight.

## Conclusion

Since based on preliminary check for 15 consecutive days, the Empirical Model is likely to be close to the truth. According to this model, the peak is expected with nearly 3300 cases per day in the fourth week of May. The number of cumulative cases will never decrease but a plateau is likely around the first week of June with only less than 100 new cases per day. The peak number of active cases is expected to be around 75,000, after which the cases with outcome (recovery or death) will be more than the number if new cases. This defines the peak requirement of the medical care facilities. The case-fatality is likely to be around 10% unless therapies such as plasma or Remedesivir succeed.

The projections are based on the trend over the previous 7 weeks and may fail if an unforeseen development occurs.

## Data Availability

Data has been taken from covid19india.org

